# A direct RT-qPCR approach to test large numbers of individuals for SARS-CoV-2

**DOI:** 10.1101/2020.06.24.20139501

**Authors:** Tomislav Maricic, Olaf Nickel, Ayinuer Aximu-Petri, Elena Essel, Marie Gansauge, Philipp Kanis, Dominik Macak, Stephan Riesenberg, Lukas Bokelmann, Hugo Zeberg, Matthias Meyer, Stephan Borte, Svante Pääbo

## Abstract

SARS-CoV-2 causes substantial morbidity and mortality in elderly and immunocompromised individuals, particularly in retirement homes, where transmission from asymptomatic staff and visitors may introduce the infection. Here we present a cheap and fast approach to detect SARS-CoV-2 in single or pooled gargle lavages (“mouthwashes”). With this approach, we test all staff at a nursing home daily over a period of three weeks in order to reduce the risk that the infection penetrates the facility. This or similar approaches could be implemented to protect hospitals, nursing homes and other institutions in this and future viral epidemics.

## Introduction

The COVID-19 pandemic has led to substantial morbidity and mortality in elderly and immunocompromised individuals, particularly in long-term care facilities (McMichael et al, 2020). Up to half of the fatalities in Europe have been observed in retirement homes (WHO, 2020), where advanced age and chronic underlying health conditions seem to predispose residents (Vishnevetsky & Levy, 2020). Therefore, it is necessary to prevent that infected personnel and visitors enter institutions where vulnerable individuals live.

However, infected individuals are often asymptomatic (Gandhi et al, 2020; Oran & Topol, 2020), resulting in a considerable rate of presymptomatic transmission of SARS-CoV-2. Furthermore, the viral load, and presumably the ability to transmit the infection, is highest at the time of symptom onset or just before (He et al, 2020). To prevent the spread of infection to institutions, one would therefore ideally test all individuals entering the facilities every time they do so. However, most commercially available viral test systems are too expensive and too laborious for such a strategy. In addition, swab kits and RNA extraction kits may run out during a pandemic (Baird, 2020).

Here, we present a safe, cheap and fast PCR-based approach to detect individuals with high levels of SARS-CoV-2 without the need for nasopharyngeal swabs or RNA extraction. We performed daily tests of all staff and residents of a local nursing home previously affected by COVID-19. We suggest that this approach could easily be scaled-up to test all staff members and visitors of retirement homes and other institutions in an entire city or region.

## Results

Nasopharyngeal swabs are commonly used to collect samples for SARS-CoV-2 testing. However, this exposes a person taking the sample to risk of infection when manipulating swabs in the nose and pharynx of potentially infectious individuals. We (in preparation) and others (Malecki et al, 2020; Saito et al, 2020) have shown that liquid washes of the oral cavity (‘gargle lavage’) can be used as a more practical, safe yet efficient alternative to nasopharyngeal swabs. This is particularly attractive for frequent sampling of asymptomatic individuals, as gargle lavages are not associated with any discomfort and can be applied to most individuals in a broad age range. We therefore focused our efforts on making such gargle lavages, commonly referred to as ‘mouthwashes’, amenable to detection of SARS-CoV-2 without isolation of viral RNA.

To this end, we investigated mouthwashes from 10 individuals who previously tested positive for SARS-CoV-2 and 10 individuals who did not. From each mouthwash, we used 200 microliter (μl) for RNA extraction in the MagNA Pure 24 System (Roche, Basel, Switzerland) and 10 μl of the resultant 50 μl RNA extract for reverse transcriptase (RT) quantitative polymerase chain reaction (qPCR) detection of SARS-CoV-2 using the LightCycler Multiplex RNA Master kit from Roche. Nine of the individuals, who on a previous occasion had tested positive, were positive in this occasion whereas none of the previously negative individuals were positive.

We then tested 1 μl of mouthwash from each of the 20 individuals using two RT-qPCR kits advertised to allow direct detection of SARS-CoV-2 from nasopharyngeal swabs: Luna Universal Probe (NEB, Ipswich, USA) and PrimeDirect (Takara, Kyoto, Japan) as well as another kit, SuperScript III with Platinum Taq (Invitrogen, Waltham, USA). The previously negative mouthwashes were negative in all cases. Among the previously positive mouthwashes, five were positive using the NEB kit whereas one and two were positive using the Takara and Invitrogen kits, respectively (Fig. 1). Although we expect the tests to be somewhat stochastic at high cycle threshold (Ct) values (the higher the Ct value, the lower the number of viral targets), the NEB Luna kit performs better with mouthwashes than the other two kits.

**Figure 1.**
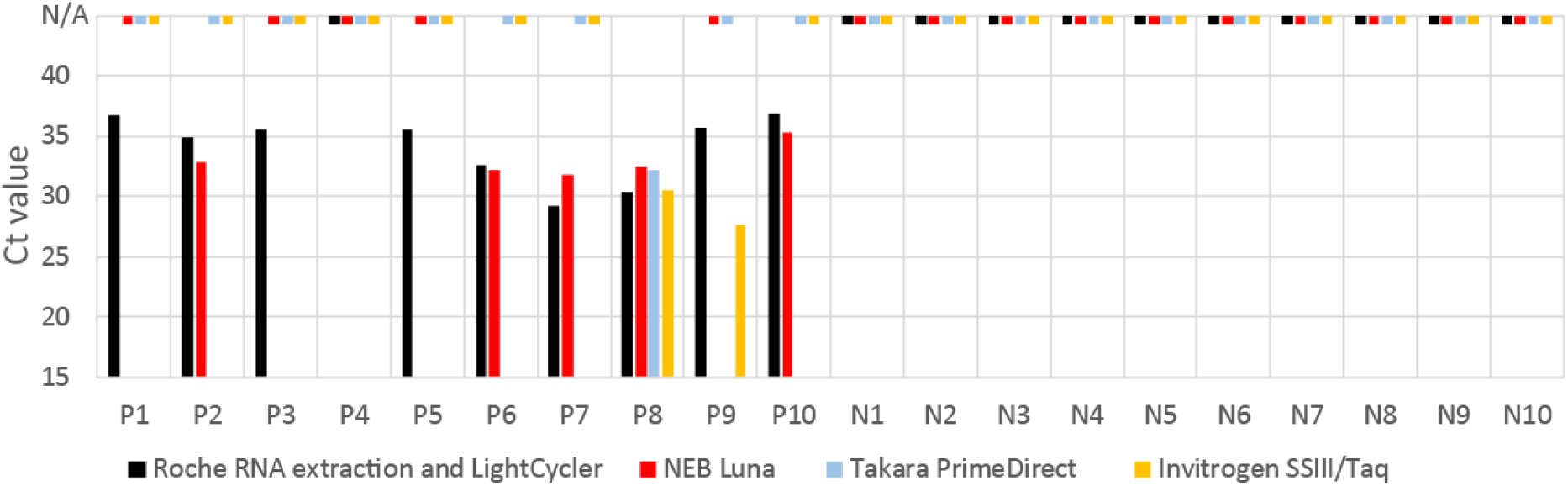
RT-qPCR of 10 previously SARS-CoV-2 positive (P) and 10 previously SARS-CoV-2 negative (N) mouthwashes. RNA was either extracted from 200 μl of mouthwashes (black bars) or 1 μl of the mouthwashes was added directly to the RT-qPCR reaction mix without prior extraction (colored bars). Dashes at N/A indicate no amplification of the viral target.

The Roche RNA extraction and qRT-PCR combination as well as the NEB kit were validated against a set of 7 reference samples distributed by “INSTAND” reference institution of the German Medical Association (Duesseldorf, Germany). These samples contain different concentrations of a heat-inactivated SARS-CoV-2 virus (sample 59, 61, 63 and 64), another coronavirus genus (sample 60 HCoV OC43, sample 65 HCoV 229E) or no virus (sample 62, supernatant of a non-infected cell culture). Fig. 2 shows that both approaches detected the SARS-CoV-2-containing samples and showed not amplification for samples not containing SARS-CoV-2.

**Figure 2.**
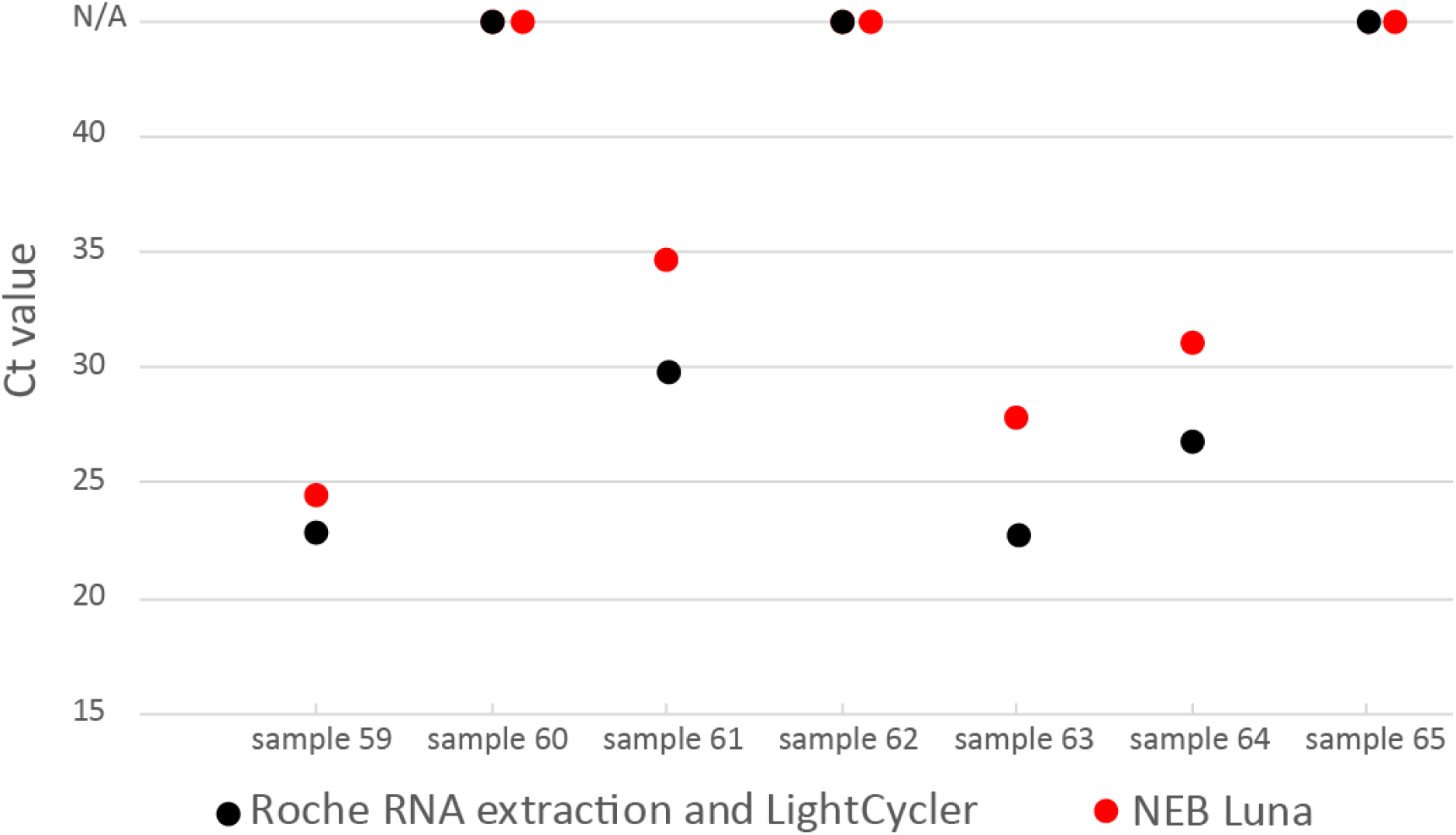
Roche RNA extraction/RT-qPCR and NEB Luna RT-qPCR of seven reference samples. Dots at N/A show no amplification of the viral target.

To systematically investigate how the NEB Luna assay performs compared to RNA extraction followed by the Roche assay for mouthwash samples, we investigated 62 gargle lavages from patients that were either negative or presented with various viral loads based on previous investigations. Fig. 3A shows that although the direct assay requires about four additional qPCR cycles on average to detect the virus, the direct assay performs well in samples where the RNA extraction-based assay detects viruses at a Ct values of ∼29 or lower, *i*.*e*. for samples that contain high viral genome counts. Among the samples positive in both methods, *i*.*e*. samples with Ct values of ∼40 or lower for the NEB assay, the results of the two methods correlate reasonably well (Fig. 3B; Pearson’s r=0.86). Notably, the extraction-free NEB assay uses 20-fold less mouthwash per assay than the Roche assay (2 μl vs. 40 μl), corresponding approximately to the average difference of the observed four PCR cycles (2^4^=16). Thus, the NEB assay *per se* is of similar sensitivity as RNA extraction followed by the Roche assay.

**Figure 3.**
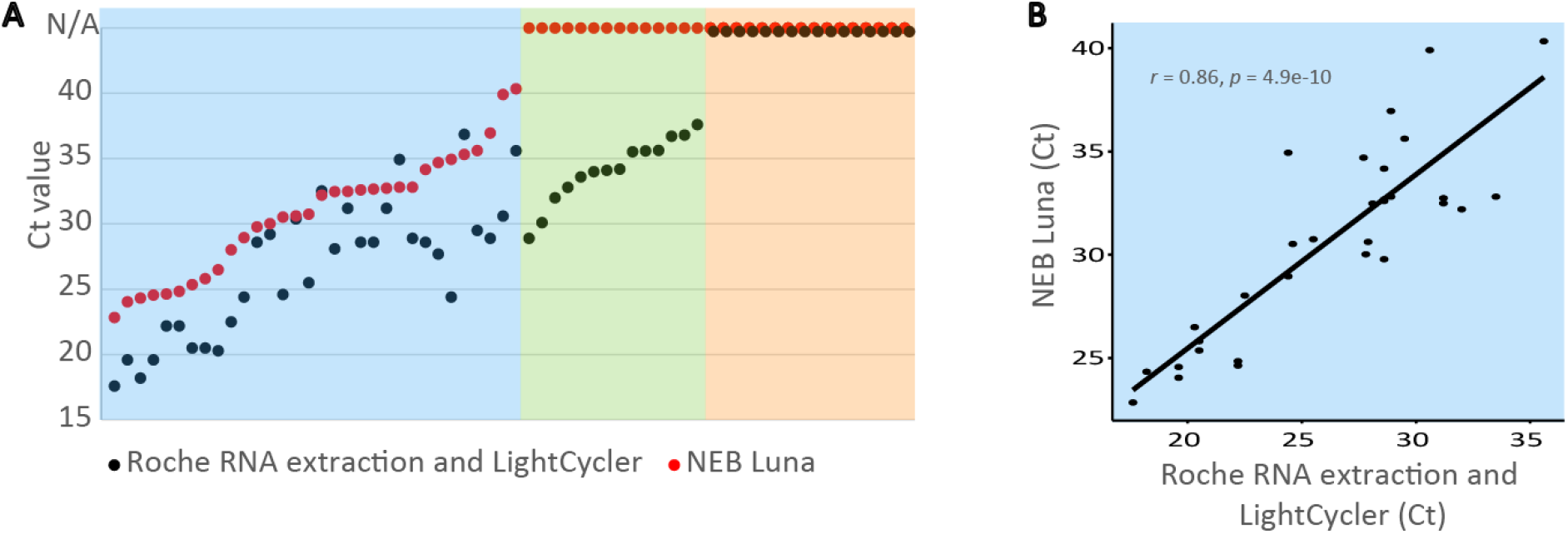
(**A**) Roche RNA extraction/RT-qPCR and NEB Luna RT-qPCR for SARS-CoV-2 detection in 62 mouthwash samples from hospital patients. The NEB method detects the virus when Ct of the Roche method is less than 29 (blue box). Dots at N/A show no amplification of the viral target. (**B**) Correlation of Ct values between direct NEB assays and RNA extraction followed by Roche assays for mouthwash samples positive with both methods.

The use of mouthwashes and direct RT-qPCR assays allow testing of large numbers of individuals. We therefore applied it to the residents and personnel of the retirement home Emmaus in Leipzig, Germany. This facility has 77 full-time residents and 88 nursing, cleaning and catering staff. In April 2020, two COVID-19 cases occurred among the residents and staff.

After appropriate informed consent, all staff working at the facility delivered a daily gargle lavage sample at the beginning or end of their working shifts. These were collected between 3 p.m. and 4 p.m. every day for 21 days and transported to the laboratory at room temperature. On weekends and holidays, the numbers of samples varied between 22 and 26 and during weekdays between 35 and 50.

We applied two testing schemes to minimize the risk that the virus could affect residents again. In the first scheme, the samples were tested individually using the direct RT-qPCR protocol and the results were evaluated and reported back to the facility by 7 p.m. To detect any inhibition that the mouthwash samples may introduce into the RT-qPCR reactions, we added a synthesized control RNA that was quantified in parallel with SARS-CoV-2 by a probe carrying a different fluorophore (Cy5) from the SARS-CoV-2 probe (FAM). For three of the 756 mouthwashes, significant inhibition was detected (Suppl. Table 1). These samples were further tested using half the mouthwash volume, which lowered the inhibition to acceptable levels (Suppl. Table 1). Nine samples were putatively positive at Ct values of 36 and higher. These were further tested by agarose electrophoresis of the PCR product and further RT-qPCR tests. In all cases, the PCR products were of unexpected sizes and renewed testing was negative (Suppl. Table 1). In summary, all 756 individual mouthwashes we tested in 21 days were negative.

In a second scheme (Fig. 4), we explored if the daily testing could be scaled up by pooling samples. To this end, we first investigated if single positive samples with Ct values between 19 and 31could be detected in pools of 26 samples. From these pools, 2 µl were used for the direct NEB assay while 200 µl were used for the RNA extraction and the Roche assay. Fig. 5 shows that in all six cases, the pools containing a mouthwash from a single infected individual were detected by the extraction-free NEB assay as well as by the standard Roche assays after RNA extraction.

**Figure 4.**
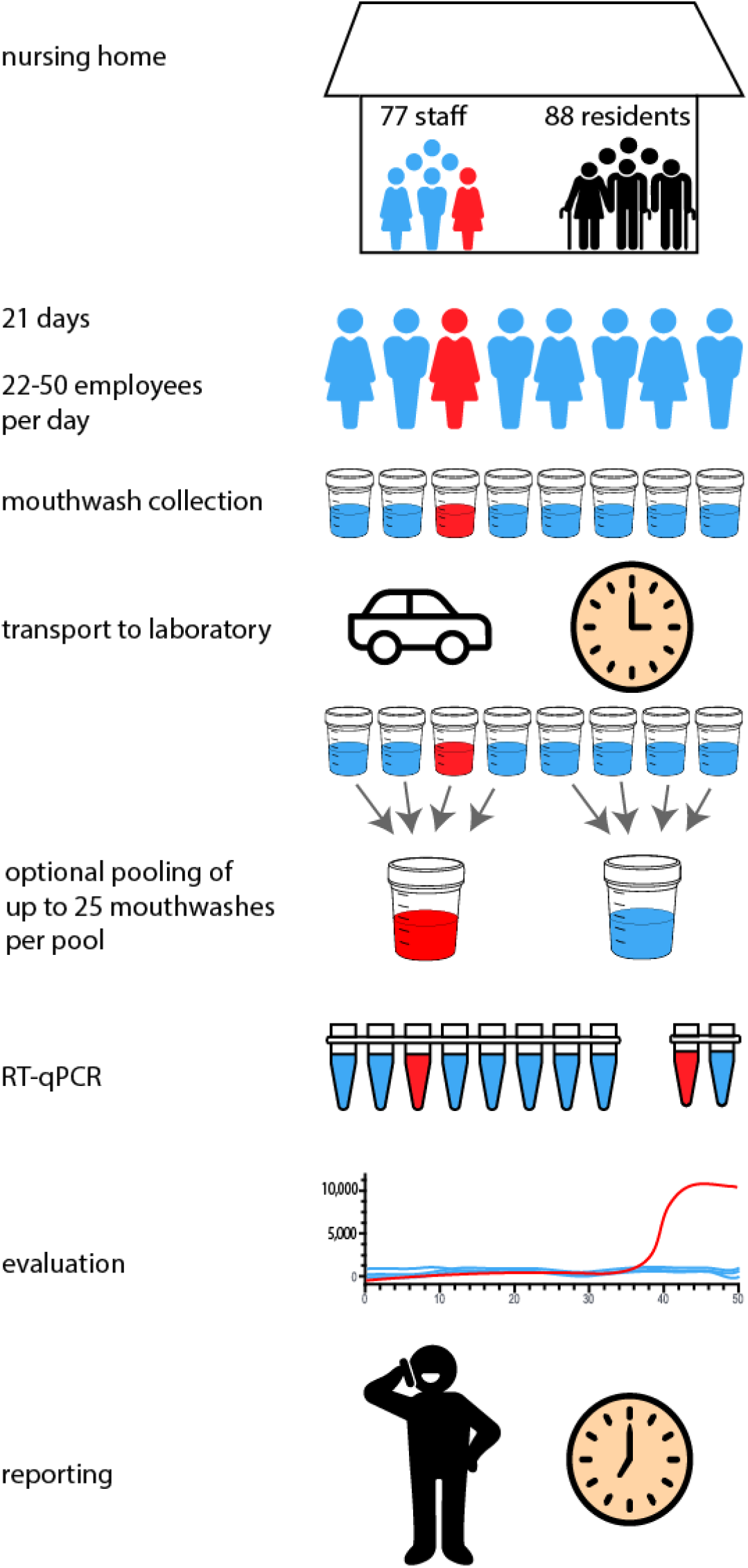
Daily testing of personnel of a nursing home with or without pooling of mouthwashes. Red color indicate a SARS-CoV-2 positive individual/sample.

**Figure 5.**
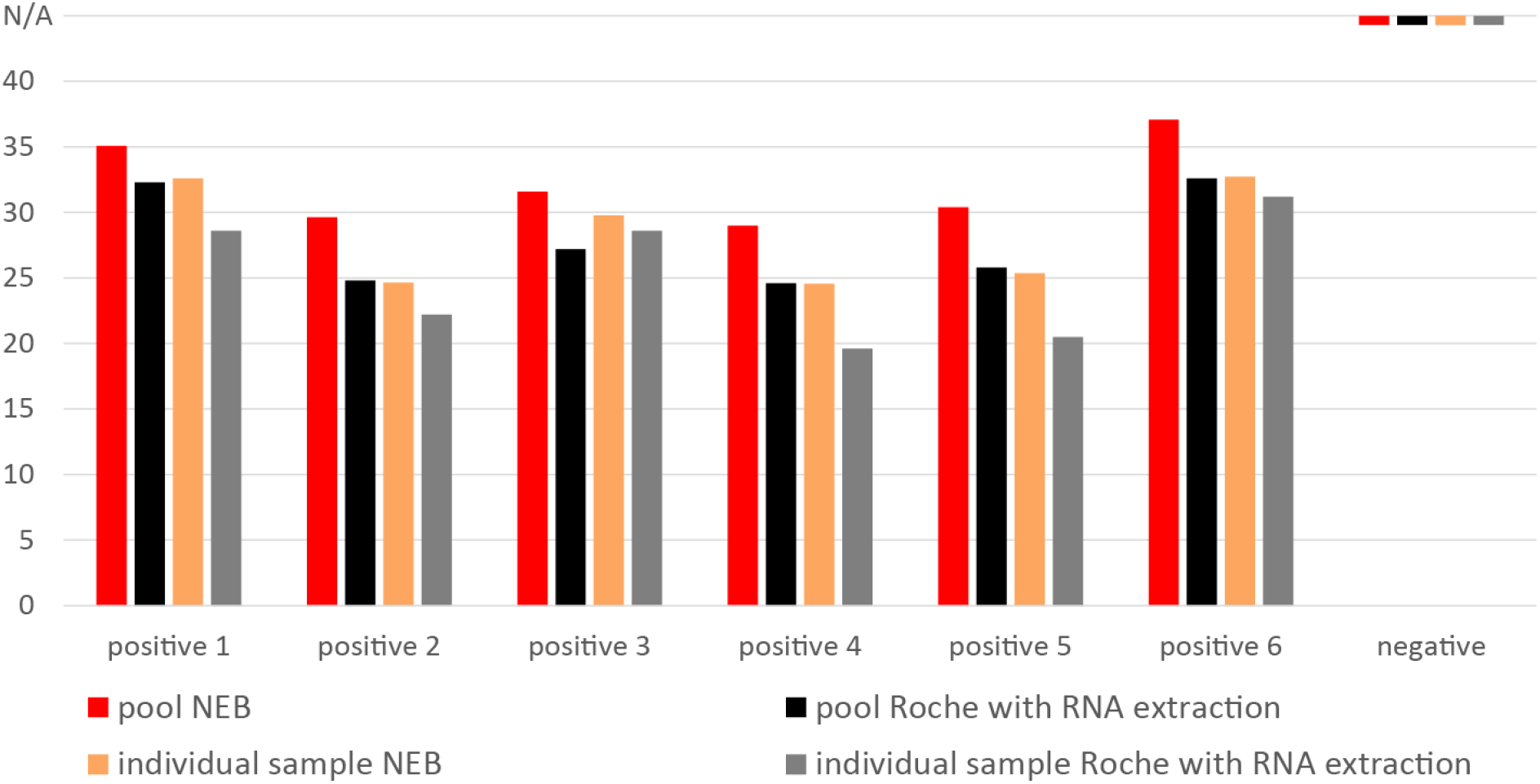
Tests of seven pools of 26 mouthwash samples, which in six cases contain one positive sample. Dashes at N/A show no amplification of the viral target.

We combined the samples from the Emmaus retirement home from each day into two pools of approximately equally many individuals and tested these pools (Fig. 4). In agreement with the individual test, none of the pools were positive. Ct values for the positive RNA control was ∼27 when no mouthwash was added and ∼28 or less for the pools, indicating that the inhibitory component seen in rare individual mouthwash samples becomes diluted and do not inhibit the reaction as much in a pool.

## Discussion

During the current SARS-CoV-2 epidemic morbidity and mortality primarily affect old or immunocompromised individuals. Thus, it is important to protect facilities where such individuals live from infections transmitted by staff or visitors. This applies not only to SARS-CoV-2 but also to other airborne communicable diseases such as influenza, RSV and adenovirus, which result in high rates of disease and death among vulnerable individuals.

To protect such individuals, it is necessary to reduce the risk of transmission from asymptomatic carriers. This is particularly the case for SARS-CoV-2, as many carriers are asymptomatic (Oran & Topol, 2020), and their infectiousness may be highest before the onset of symptoms (He et al, 2020; Wolfel et al, 2020). It is therefore desirable to test asymptomatic individuals that come into contact with vulnerable individuals as frequently as possible. The protocol described here enables large-scale high-throughput screening at a reagent costs of about 2-4 Euros per sample (in 2020 NEB Luna kit cost ∼0.6 Euros per sample and primer/probes cost 1.5-3.5 Euros depending on the provider).

Three factors make this approach suitable for implementation on a large scale. Firstly, gargle washes rather than nasopharyngeal swabs have the advantage that they can be collected by the individuals themselves and do not expose any personnel performing sampling to risk of infection. They are also not associated with any discomfort for the person being tested so that they can easily be performed on a daily basis. Secondly, since the protocol does not require isolation of RNA, it reduces the need for equipment, reagents, labor, time and money allowing larger numbers of tests to be performed. Thirdly, pooling of samples allows large numbers of individuals to be tested. A practical approach might be to divide the staff at each institution into two pools to allow one group to return to work even if the other group would turn out to contain one or more positive individuals. Using this approach, about 45 institutions of the size of the retirement home tested here could be analyzed on a single, 96-well microtiter plate in about an hour and half (∼3,700 individuals, ∼300 Euros total reagent cost).

In principle, it would be preferable to test staff members before starting their work shifts and come into contact with residents. However, including transport and evaluation, between two and five hours are needed before results can be reported back to the facility. Thus, it is more practical to perform the tests in the evening. If one individual, or one pool of samples, is found to be positive, that individual or group of individuals can then be prevented from working the next day, and, in the case of pooled samples, be individually tested. In the future, other methods such as reverse transcriptional loop-mediated isothermal amplification (RT-LAMP) (Yu et al, 2020) may be developed to a point where they can performed reliably locally at facilities at risk such that staff can be tested before they start their daily work.

The RNA isolation-free assays are slightly less sensitive than the standard assay due to the smaller volume of mouthwash that can be used in the assay. However, there is evidence that only individuals with a high viral load (a Cq value of 30 or lower) are likely to transmit the infection (NCID, 2020; Wolfel et al, 2020; Zhou et al, 2020). Thus, we suggest that the advantage of daily testing greatly outweighs the lower sensitivity in the low-copy range.

We note that if a rapid and inexpensive testing scheme like the one presented here is implemented, personal visits to individuals at risk could be allowed with large social benefits. For example, if visitors were tested in the morning they could, if negative, visit in the afternoon. We furthermore note that simplified testing could also be applied to asymptomatic travelers to slow the spread of infections - while still allowing global travel. Finally, we can foresee that with daily testing of staff members and visitors of retirement homes, hospitals, prisons and critical infrastructure, society may be able to sustain function with minimal restrictions in cases where a virus poses little threat to young and healthy individuals as is the case for the current SARS-CoV-2 and seasonal influenza outbreaks.

Future evolution of SARS-CoV-2 or other viruses could result in strains causing an increased major morbidity also in the general population. Obviously, large-scale applications of simplified tests of pooled samples as described here are likely to be extremely valuable to contain also such situations.

## Materials and Methods

All individuals included in this study were asked for their voluntary assistance to participate. Each individual gave written informed consent before entry into the study. The study was approved by the Ethics Committee of the Saxonian medical chamber (EK-allg-37/10-1). All procedures utilized in this study are in agreement with the 1975 Declaration of Helsinki.

### Gargle lavages (‘mouthwashes’)

For mouthwashes, 10 mL of sterile water was pre-filled into sterile scaled urine cups. The individuals to be tested took the full volume of water into the mouth and gargled for 10 seconds. Subsequently, the mouthwash was spitted back into the cup and firmly closed with a watertight lid and stored at room temperature for up to 4-8 hours.We also analyzed anonymized leftover material from hospitalized COVID-19 patients. In these cases, mouthwashes had been stored frozen below −80°C for 1-2 months.

### Primers and probes for RT-qPCR

All primers, probes and controls were from Tib-Molbiol (Berlin, Germany). For SARS-CoV-2 detection, the E-gene was detected with a FAM probe (Cat.-No. 53-0776-96). For inhibition tests, EAV RNA Extraction Control 660 with Cy5 probe (Cat.-No. 66-0909-96) was used.

### Roche RNA extraction and RT-qPCR

Individual mouthwashes were vortexed and 200 µl were used for RNA extraction with the MagNA Pure 24 System from Roche based on the manufacturer’s protocol. RNA was eluted in 50 µl of which 10 µl was used in the RT-qPCR LightCycler Multiplex RNA Master kit with 4.9 µl water, 0.5 µl E-gene primer and probe reagent mix, 0.5 µl EAV control primer and probe reagent mix, 4 µl RT-PCR reaction mix and 0.1 µl RT-enzyme solution. The RT reaction was performed at 55°C for 5 minutes. qPCR included a denaturation step at 95°C for 5 minutes, followed by 45 cycles with 5 seconds at 95°C, 15 second at 60°C and 15 seconds at 72°C. RT-qPCR reaction was performed on a and analyzed with Roche LightCycler 480 Software.

### Direct RT-qPCR

Mouthwashes were vortexed and either 1 or 2 µl were added into a reaction tube containing pre-assembled reagents. RT-qPCR reaction was run on BioRad CFX96 Real-Time system and results analyzed with Bio-Rad CFX Manager software where the Ct determination mode was set to “regression”.

### Takara PrimeDirect Probe One-step RT-qPCR

Twenty five µl reactions contained: 12.5 µl prime direct mix, 0.5 µl E-gene primer and probe reagent mix, 1 µl mouthwash and 11 µl water. The reaction was incubated at 90°C for 3 minutes, 60 °C for 5 minutes followed by 50 cycles of 95 °C for 5 seconds and 55 °C for 30 seconds.

### Invitrogen SuperScript III with Platinum Taq One-Step RT-qPCR

Twenty five µl reaction contained: 12.5 µl rxn mix, 0.5 µl E-gene primer and probe reagent mix, 0.4 µl MgSO4, 1 µl enzyme mix, 1 µl mouthwash and 9.6 µl water. The reaction was incubated at 60 °C for 5 minutes, 55 °C for 5 minutes, 95 °C for 2 minutes followed by 50 cycles of 94 °C for 5 seconds, 60 °C for 30 seconds and 68 °C for 10 seconds.

### NEB Luna Universal Probe One-Step RE-qPCR kit

Twenty five µl reaction contained: 12.5 µl Reaction MasterMix, 1.25 µl EnzymeMix, 0.5 µl E-gene primer and probe reagent mix, 0.5 µl EAV control primer and probe reagent mix, 0.5 µl EAV control (pellet eluted in 1,200 µl TE buffer), 1-2 µl mouthwash, and 8.75-9.75 µl water. For Fig.1, 1 µl of mouthwash was used, for later experiments 2 µl to allow for more accurate pipetting. Mastermix can be stored in the fridge up to a week. RT-qPCR program for the NEB reaction was: 55 °C for 4 minutes, 60 °C for 4 minutes, 55 °C for 2 minutes, 95 °C for 1 minute followed by 50 cycles of 95 °C for 10 seconds and 60 °C for 30 seconds.

## Data Availability

Data is available in figures and the supplementary table. Data that is not there will be made available upon request.

## Acknowledgements

The authors thank the management of Diakonische Dienste Leipzig, the personnel and residents of Altenpflegeheim Emmaus in Leipzig, Germany, for their cooperation. Furthermore, we recognize the efforts and helpful instructions contributed by Nils Lahl from the public health department of the City of Leipzig. We particularly thank Jeannine Dulz, Katrin Grabietz, Jeannette Hofmann, Michelle Karius, Sandra Reinhardt, Kerstin Rolle, Katharina Wald, Kristina Winter for assisting and performing reference and confirmatory testing at the Department of Laboratory Medicine at Hospital St Georg Leipzig. Funding was provided by the Max Planck Society and the NOMIS foundation.

## Notes

### Competing Interest Statement

The authors have declared no competing interest.

